# Development and Clinical Application of a Deep Learning-Based Endometrial Cancer Cytology Supporting Model

**DOI:** 10.1101/2025.05.21.25327411

**Authors:** Ichito Shimokawa, Mika Terasaki, Shun Tanaka, Etsuko Toda, Shoichiro Takakuma, Yusuke Kajimoto, Shinobu Kunugi, Akira Shimizu, Yasuhiro Terasaki

## Abstract

**Background:** The global rise in endometrial cancer, including in Japan, and the shortage of pathologists and cytotechnologists has increased the diagnostic burden, emphasizing the need for AI-based diagnostic support model using deep learning. This study aims to advance an existing AI-supported endometrial cytology model for clinical application.

**Methods:** We compared two datasets—one annotated for both benign and malignant clusters, and one for malignant only—using YOLOv5x and YOLOv7 models evaluated by mAP. We also assessed the correlation between AI diagnostic accuracy and the level of difficulty perceived by human diagnosticians using the Two One-Sided Tests (TOST) procedure. Additionally, we applied Grad-CAM to visualize and enhance the interpretability of the AI model’s decision-making process.

**Results:** The YOLOv5x model with both benign and malignant annotations achieved the highest malignant mAP at 0.798 compared to Yolov7. The TOST analysis showed no significant difference in perceived diagnostic difficulty between cases that were correctly and incorrectly diagnosed by the AI model, indicating consistent AI accuracy regardless of case difficulty. Grad-CAM visualizations clarified the AI model’s decision-making basis; in some cases, the model appeared to focus on regions different from those typically attended to by human diagnosticians.

**Conclusion:** The AI support model showed high and consistent accuracy in endometrial cytology, regardless of diagnostic difficulty as perceived by human diagnosticians. Grad-CAM visualizations revealed diagnostic patterns, with AI occasionally focusing on regions different from those emphasized by human diagnosticians. This study advances the real-time, microscope-integrated AI system toward clinical application.

## 1. Introduction

Endometrial cancer is a gynecological malignancy with increasing incidence rates worldwide, ^1^ particularly in developed countries, where it is the most prevalent. Most cases occur in the postmenopausal period, with a mean age of 60 years at presentation. Early detection and appropriate treatment have a profound impact on patient survival and quality of life. For example, the five-year survival rate for stage I endometrial cancer exceeds 90% but drops to approximately 70% for stage III disease. ^2^ This emphasizes the importance of early diagnosis and highlights the need for rapid and accurate diagnostic methods.

Endometrial cytology is a crucial diagnostic method for early detection of endometrial cancer.^3^ Since abnormal uterine bleeding is often the first symptom in many cases, it is recommended to perform cytology promptly when such bleeding is observed. ^4^ This test is widely used because it can be easily performed in an outpatient setting with minimal burden on the patient. In endometrial cytology, cells are directly collected from the endometrium using a specialized instrument, and these cells are then examined under a microscope to detect the presence of cancer cells at an early stage. However, this test presents certain diagnostic challenges. The morphology of cells can easily change under the influence of hormones, ^5^ and the thickness and overlap of cell clusters can make diagnosis difficult, requiring advanced expertise and technical skills.

Deep learning, a subset of AI, has gained significant attention in recent years for its potential in various fields, particularly in medical image analysis, for example, for the detection of lung cancer using chest X-ray images, ^6^ of colon cancer ^7^ through endoscopic live images and for diagnosing diabetic retinopathy ^8^ through retinal images. The potential for AI to streamline workflow and support decision-making extends beyond specific diseases, potentially benefiting a wide range of clinical applications from early screening to complex case management. Deep learning has been applied in various fields within pathology, ^9^ however, applying it specifically to endometrial cytology presents unique challenges. One such challenge is obtaining clear digital images of tissue samples, known as Whole Slide Imaging (WSI). ^10^ In endometrial histology, WSI has been successfully applied to AI models for diagnostic purposes, however, cytology faces additional challenges due to the thickness and overlap of cell clusters^11^. WSI involves scanning entire glass slides to create high-resolution digital images but achieving consistent focus across thick or overlapping cell clusters can be difficult when dealing with directly smeared cells on the glass slide, as opposed to uniformly thin sections from formalin-fixed, paraffin-embedded specimens. This inherent complexity in cytology has slowed the development of AI models in endometrial cytology.

In response to these challenges for obtaining digital images from endometrial cytological slides, we have developed an AI-assisted system for the detection of endometrial cancer cells that operates in real-time under a microscope without requiring WSI scans in our previous research.^12^ We used a pre-existing deep learning model known as YOLOv5x (You Only Look Once version 5x), which is specifically designed for rapid object detection, and customized the model to suit our specific dataset. This system is designed to seamlessly integrate into existing microscope workflows, allowing clinicians to verify AI-generated assessments as they view the slides. Our earlier studies demonstrated that this system could significantly reduce diagnostic time across evaluators with varying levels of experience, from skilled pathologists to medical students, while also alleviating the workload on diagnosticians and improving diagnostic accuracy.

Our previous research demonstrated the usefulness of an AI-assisted system in endometrial cytology, but it also revealed several significant limitations. Training an AI model typically requires a large amount of labeled data to teach it what to recognize. For instance, to enable the AI to differentiate between cancerous and normal cells, regions containing cancer cells are labeled as “malignant”, while the labeling strategy for normal cell clusters is often not well defined. Moreover, it remains unclear which annotation approach yields the best performance. This uncertainty affects the reliability of AI decisions and warrants further investigation.

Next, we chose to use the YOLOv5x model. This model is well-suited for our task because it can rapidly identify specific areas in microscope images where cancerous cells might be present. However, we have not yet compared the performance of YOLOv5x with other AI models to see if it is truly the best option for this type of work. Evaluating and comparing different models is important because it helps us ensure that we are using the most effective tool for accurately detecting cancer cells. This is an issue that warrants further investigation in future research.

Many studies on medical AI models emphasize quantitative metrics such as object or abnormality detection accuracy. While such metrics are useful for evaluating technical performance, they often fail to capture the true clinical value of AI systems. To address this limitation, it has been suggested that comparing diagnostic performance between AI-assisted and unassisted workflows is essential for assessing clinical applicability.^13^ In our previous research, we responded to this need by conducting a comparative analysis of diagnostic performance with and without AI support. ^12^ However, we did not examine the relationship between diagnosticians’ perceived case difficulty and whether the AI model made a correct or incorrect prediction. This omission may have limited our understanding of the model’s practical utility in real-world settings, and ignoring this aspect could lead to a misleading interpretation of its usefulness in clinical practice.

Additionally, AI models that use deep learning face what is known as the “black box problem”.^14^ Deep learning-based models consist of multiple computational layers that automatically extract hierarchical features from the input data. While this allows for high performance, the depth and complexity of these layers make it difficult to understand the decision-making process. This opacity is referred to as the “black box problem,” highlighting the challenge of interpreting how the AI model arrives at its diagnostic conclusions. If we cannot determine which aspects the AI focused on and why it reached a particular conclusion, it raises concerns about the reliability of the diagnosis. To address this issue, it is essential to develop “explainable AI (XAI)” that can clarify how the AI makes its decisions.

Using a technique called Grad-CAM (Gradient-weighted Class Activation Mapping), which visualizes the regions that most strongly influence the model’s output by utilizing the gradients of the final convolutional layer, we can visualize which parts of an image the AI focused on during a specific diagnosis .^15^ This allows us to visually confirm how the AI arrived at its decision, making the rationale behind the diagnosis more understandable. It is crucial to utilize such techniques to clarify the basis on which the AI diagnoses benign and malignant cell clusters.

To address the limitations identified in our previous research, this research focuses on four key objectives:

1. **Annotation Strategy**: We will investigate whether labeling both benign and malignant cell clusters or labeling solely on malignant clusters provides better diagnostic accuracy. This will help us determine the most effective annotation strategy for training AI models.
2. **Model Comparison**: We will compare the performance of the YOLOv5x model with other AI models to evaluate which is the most effective for detecting cancer cells in endometrial cytology. This comparison is crucial to ensure we are using the best possible tool.
3. **Integrating the AI Model into Routine Diagnostic Practice**: We will examine the correlation between the perceived difficulty of diagnosis by pathologists and cytotechnologists and the accuracy of the AI model. This analysis will help us align AI models with the practical challenges faced in clinical settings.
4. **Explainability and Transparency**: We will explore the use of techniques such as Grad-CAM to enhance the explainability of AI models. By visualizing the AI’s decision-making process, we aim to improve the transparency and trustworthiness of AI-assisted diagnoses in clinical practice.

These objectives will guide the refinement of our AI-assisted system for endometrial cytology, enhancing its accuracy, reliability, and clinical suitability. Moreover, these areas of investigation hold significance beyond this model, supporting the broader development of AI applications across medical imaging and healthcare innovation.

## 2. Method

### 2.1. Ethics

Ethical approval for this study was obtained by the Institutional Review Board of Nippon Medical School (approval number: 23K08900).

### 2.2 Patient Data Collection

Patient selection and cytology preparation followed the methodology described in our previous study^12^, which detailed the collection and classification of endometrial cytology cases.

From April 2017 to March 2023, a total of 96 cytology slides were collected at Nippon Medical School Hospital. Among these, 47 cases were diagnosed with “Malignant” endometrial cancer, while the remaining 49 cases were classified as “Benign”, including non-malignant endometrial conditions such as leiomyoma. Table 1 outlines the case distribution and provides the median age for each category. All diagnoses were pathologically confirmed with hysterectomy specimens. The endometrial cytology samples were prepared by smearing and stained using the Papanicolaou method.

**Table 1.**
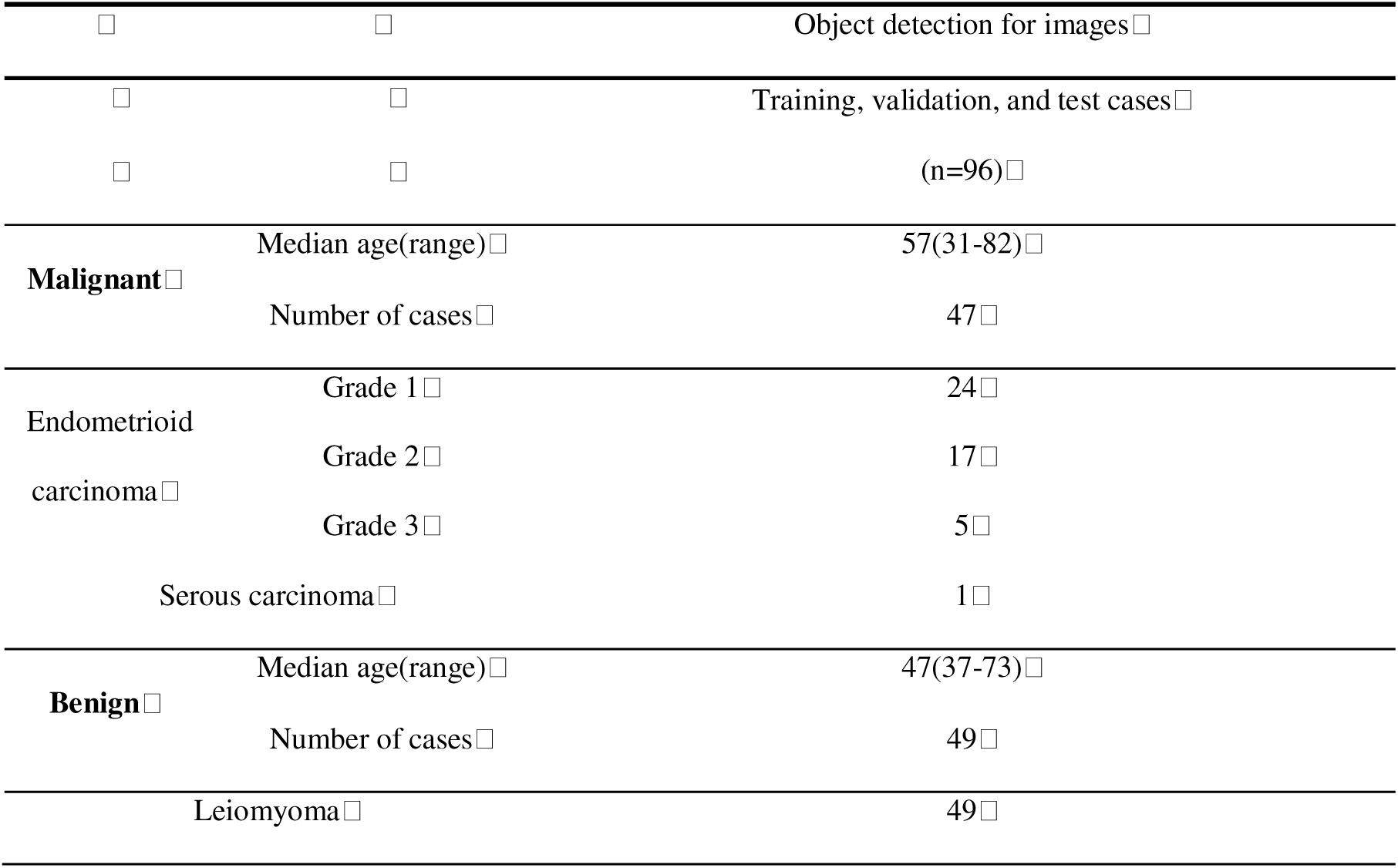
Case Presentation.

### 2.3 Acquisition of Digital Images Using a Smartphone Diagnostic Device

Digital images were acquired using the smartphone-based system as described in our earlier work.^12^ Detailed procedures, including device specifications and imaging conditions, are provided in that report. Digital images were obtained using a smartphone-enabled diagnostic setup. An iPhone SE (Apple Inc., Cupertino, CA, USA) was attached to an Olympus BX53 microscope (EVIDENT, Olympus, Tokyo, Japan) via a specialized adapter (i-NTER LENS; Micronet Co., Ltd., Kawaguchi, Saitama, Japan). The images were captured at a resolution of 4032×3024 pixels. The focus was manually adjusted during direct observation of the cytology slides, and imaging was performed with a 20x objective lens. For malignant cases, images were centered on clusters of abnormal cells identified by a gynecologic pathologist. In benign cases, normal cell clusters were randomly selected for imaging.

### 2.4 Creation of Image Datasets

For deep learning model training, a total of 1536 malignant cell cluster images and 2278 benign cell cluster images were divided into training data, validation data, and test data in an 8:1:1 ratio. The breakdown is shown in Table 2.

**Table 2.** Number of malignant and benign images for each dataset.lJ.

| Dataset | Training Data | Validation Data | Test Data |
| --- | --- | --- | --- |
| Malignant images | 1234 | 154 | 148 |
| Benign images | 1819 | 231 | 228 |
| total | 3053 | 385 | 376 |

### 2.5 Image Preprocessing and Annotation

The collected images, originally 4032×3024 pixels, were resized to 800×600 pixels for training. Annotation was performed using the Python-based software LabelImg (version 1.8.6). Two datasets were created: one where only malignant cell clusters were annotated, treating benign cell clusters as background, and another where both malignant and benign cell clusters were annotated. This process is illustrated in Figure 1.

**Figure 1.**
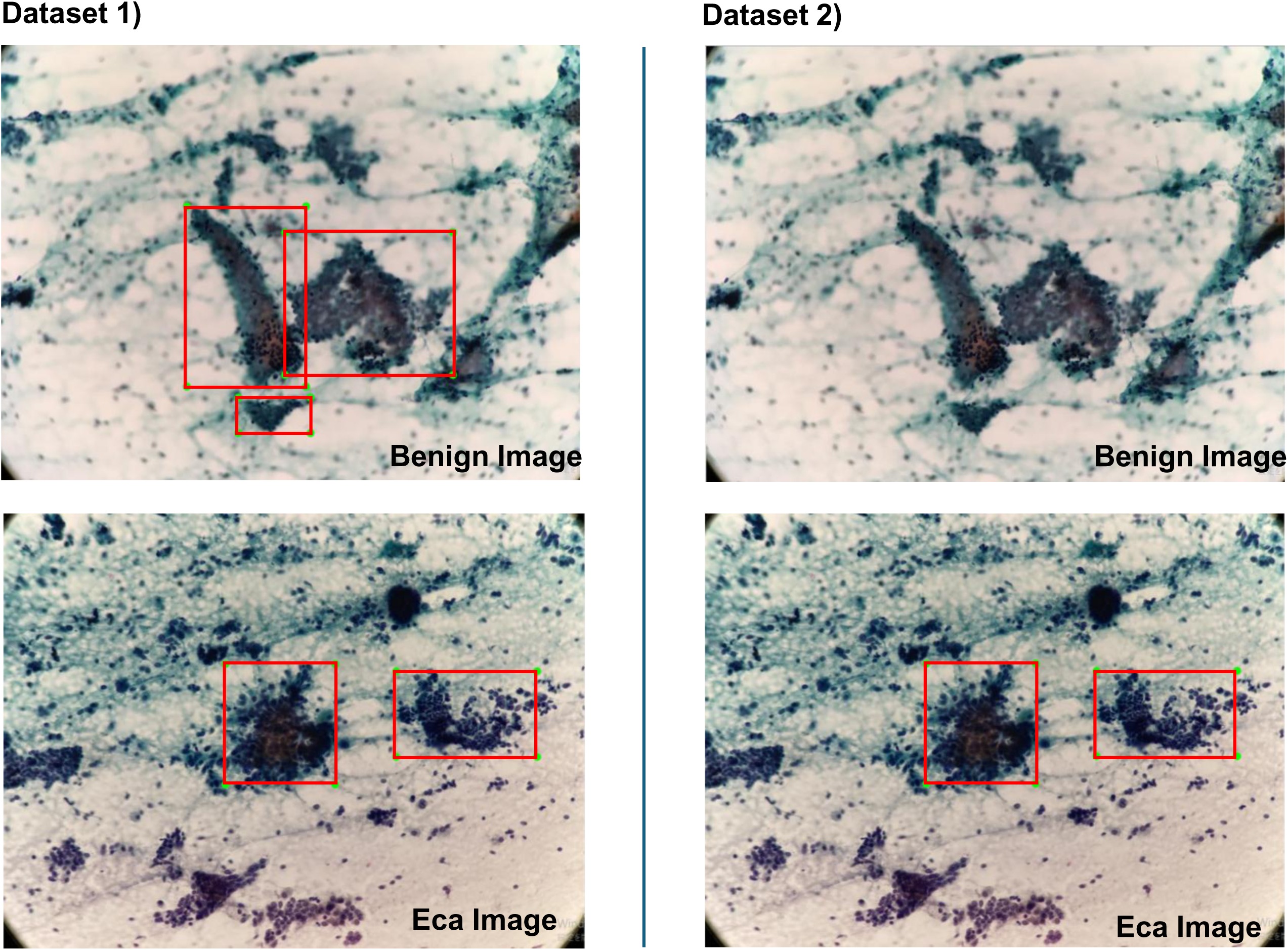
The process of creating datasets. In Dataset 1, both benign and malignant cell clusters are annotated. In Dataset 2, only malignant cell clusters are annotated.

### 2.6 Models and Their Architecture

The YOLO series is widely used in medical image analysis for object detection tasks, owing to its high accuracy and real-time performance.^16^ Specifically, YOLOv5x and YOLOv7 are suited for clinical environments that require high precision and fast processing capabilities for real-time diagnosis.

In medical image analysis, rapid and accurate diagnosis are essential, and the high frame rate capability of YOLO Series is crucial.^17^ These models efficiently process high-resolution images, reducing diagnostic delays. Moreover, YOLOv5x and YOLOv7 offer advanced feature extraction to detect complex and subtle abnormalities, which is crucial for medical images. This enables the precise detection of small and ambiguous boundary lesions that might otherwise go unnoticed. Therefore, this study compares the accuracy of YOLOv5x and YOLOv7.

YOLOv5x provides high performance and flexibility in object detection. Its main features include a three-layer structure (backbone, neck, and head). This structure enables YOLOv5x to build robust models for a wide range of applications. It excels in real-time, high-precision object detection required in medical image analysis. Additionally, YOLOv5x achieves a lightweight model design and faster processing speed, making it effective for use on edge devices and in resource-limited environments.

YOLOv7, an improved version of the YOLOv5x series, utilizes Extended Efficient Layer Aggregation Networks (E-ELAN) to enhance feature learning ability without changing the gradient propagation path and to improve parameter and computational efficiency.^18^

The selected YOLO models were pre-trained on the Microsoft COCO dataset and fine-tuned using our medical dataset through transfer learning. By transfer learning these pre-trained models, the YOLO series can adapt effectively to the specific challenges of cytology images in medical image analysis.

### 2.7 Deep Learning Model Training

We adopted two pre-trained models, YOLOv5x and YOLOv7, and experiments were conducted over 100 epochs with a batch size of 4, and hyperparameters were manually tuned.

The primary evaluation metric used to assess AI model performance in this study was mean Average Precision (mAP). mAP is widely used to assess model accuracy, especially in classification and object detection tasks, by calculating the average precision (AP) for each class and taking their mean. AP evaluates the relationship between precision and recall, providing a comprehensive accuracy metric independent of specific thresholds.

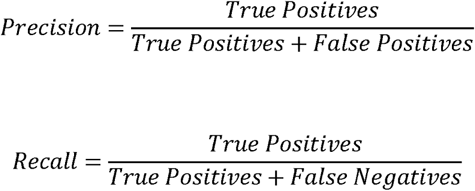

AP is obtained by plotting the precision-recall curve and calculating its Area Under the Curve (AUC). Specifically, precision and recall are calculated for different thresholds for each class, and the area under the precision-recall curve is computed as AP. The mAP is obtained by averaging the AP for all classes.

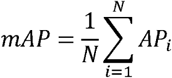

where N is the total number of classes, and *AP_i_* is the average precision for class *i.* mAP is a comprehensive metric that evaluates a model’s prediction accuracy across all classes. It is especially useful in multi-class classification and object detection tasks. High mAP values indicate that the model balances high precision and recall, demonstrating its utility as a reliable diagnostic support tool.

### 2.8 Execution Environment

A notebook PC, “Iiyama Sense 15F161,” with an Intel Core i7 CPU and an NVIDIA GeForce RTX3060 GPU was utilized. The software environment consisted of the Anaconda distribution (version 2022.10), Python 3.9.16 as the programming language, and PyTorch 1.13.1 as the deep learning framework.

### 2.9 Comparison with Perceived Diagnostic Difficulty by Cytotechnologists and Pathologists

As shown in Table 2, the created dataset was divided into training, validation, and test data in an 8:1:1 ratio.

During the model training of YOLOv5x and YOLOv7, training data was used for training, and validation data was used for inference regarding model accuracy metrics. Finally, test data was read by YOLOv5x to investigate the correlation between the diagnosticians’ perceived difficulty and the AI model’s accuracy in determining malignant and benign cases. This was because the YOLOv5x model produced the best results as shown later. From the test set, 20 images were extracted—five each from the following four categories: (1) benign cases misclassified as malignant, (2) benign cases correctly classified, (3) malignant cases misclassified as benign, and (4) malignant cases correctly classified. These were grouped into two sets: 10 images correctly classified by the AI and 10 misclassified.

Eight cytotechnologists and three pathologists who did not participate in image capture or model creation evaluated the difficulty of each image on a scale of 1 (easiest) to 5 (most difficult). The evaluators were not informed of whether the images were benign or malignant. This process is summarized in the flowchart in Figure 2.

**Figure 2.**
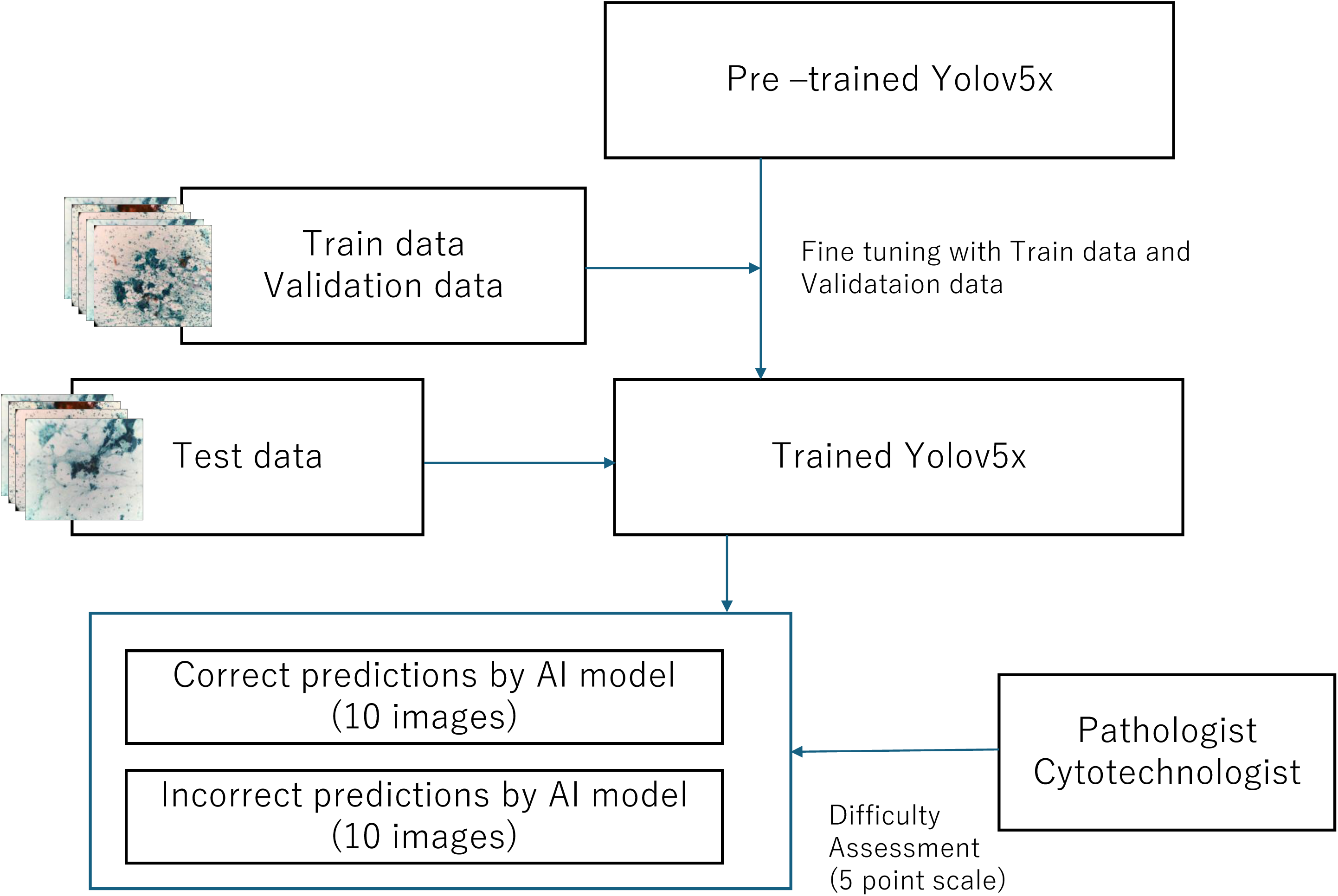
Simple flowchart of Model Training, test, and Evaluation of Diagnostic Difficulty Perceived by Pathologists and Cytotechnologists

### 2.10 Calculation of Mean Difficulty Scores and Box Plot Creation

To quantify the perceived difficulty of each image, we calculated the mean difficulty scores provided by the diagnosticians for both correctly and incorrectly classified image groups. The results were visualized using box plots, which illustrate the central tendency and variability of the difficulty scores in each category.

This analysis aimed to investigate whether cases misclassified by the AI were systematically associated with higher perceived difficulty by human diagnosticians.

### 2.11 Statical analysis

To examine whether the difficulty scores for correctly classified and misclassified images by AI are equivalent, we conducted an equivalence test (Two One-Sided Tests, TOST).^19^ The equivalence range was set at ±0.5. The standard error (SE) of the sample difference is calculated as follows:

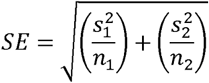

where *s_1_* and *s_2_* are the standard deviations of each group. The t-values for the lower and upper bounds are calculated using:

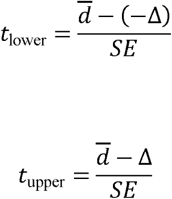

The cumulative distribution function (CDF) is then used to determine the p-values. If the p-values are below 0.05, the two groups are considered statistically equivalent within the specified range.

### 2.12 Visualization of Diagnostic Basis for Benign and Malignant Diagnosis by YOLOv5x

In this study, a model combining YOLOv5x and Grad-CAM (Gradient-weighted Class Activation Mapping) was used to visualize the diagnostic basis of the AI model. Grad-CAM is a technique that visualizes the areas of interest in the model for a specific input image, helping to understand which parts of the image the model focuses on. Specifically, Grad-CAM was applied to the final convolutional layer of the YOLOv5x model to reveal the regions of the image that the model focused on when diagnosed as benign or malignant. The images generated by this technique highlight the model’s reasoning for specific diagnostic outcomes. The heatmaps created by Grad-CAM emphasize key regions in the image, clarifying the features the model relied on to make its diagnosis. For example, if the model diagnosed malignancy, the regions contributing to this decision, such as abnormal cell clusters, are shown in high-intensity colors like red or yellow. Conversely, if the model diagnosed benignity, normal tissue structures or non-abnormal cell clusters are highlighted.

## 3. Results

### 3.1 Training and Accuracy of the Benign and Malignant Mixed Models

Two types of training models were developed using both YOLOv5x and YOLOv7: one where both malignant and benign cell clusters were annotated, and another where only malignant cell clusters were annotated. The accuracy metrics for these models are as follows:

- **YOLOv5x (mixed annotation)**: A malignant mAP of 0.798 and a benign mAP of 0.676.
- **YOLOv7 (mixed annotation)**: A malignant mAP of 0.781 and a benign mAP of 0.663.
- **YOLOv5x (malignant-only annotation)**: A malignant mAP = 0.769.
- **YOLOv7 (malignant-only annotation)**: A malignant mAP = 0.647.

These results are summarized in Table 3.

**Table 3.** The mAP of models of YOLOv5x and YOLOv7.

|  |  | Benign and Eca annot | Only Eca annot |
| --- | --- | --- | --- |
| YOLOv5x | Eca | **0.798** | 0.769 |
|  | Benign | 0.676 |  |
| YOLOv7 | Eca | 0.781 | 0.647 |
|  | Benign | 0.663 |  |

Abbreviation: mAP50, mean Average Precision (IOU threshold = 0.5) , Eca means endometrial cancer.

In this study, the YOLOv5x model with mixed benign and malignant annotations achieved the highest accuracy, with a mean Average Precision (mAP) of 0.798 in detecting malignant clusters. YOLOv5x outperformed YOLOv7 in both “Benign and Malignant” (mAP 0.798 vs. 0.781) and “Malignant-only” (mAP 0.798 vs. 0.647) annotation settings, demonstrating its superior performance in identifying malignant clusters. Figure 3 illustrates representative detection outputs from each model.

**Figure 3.**
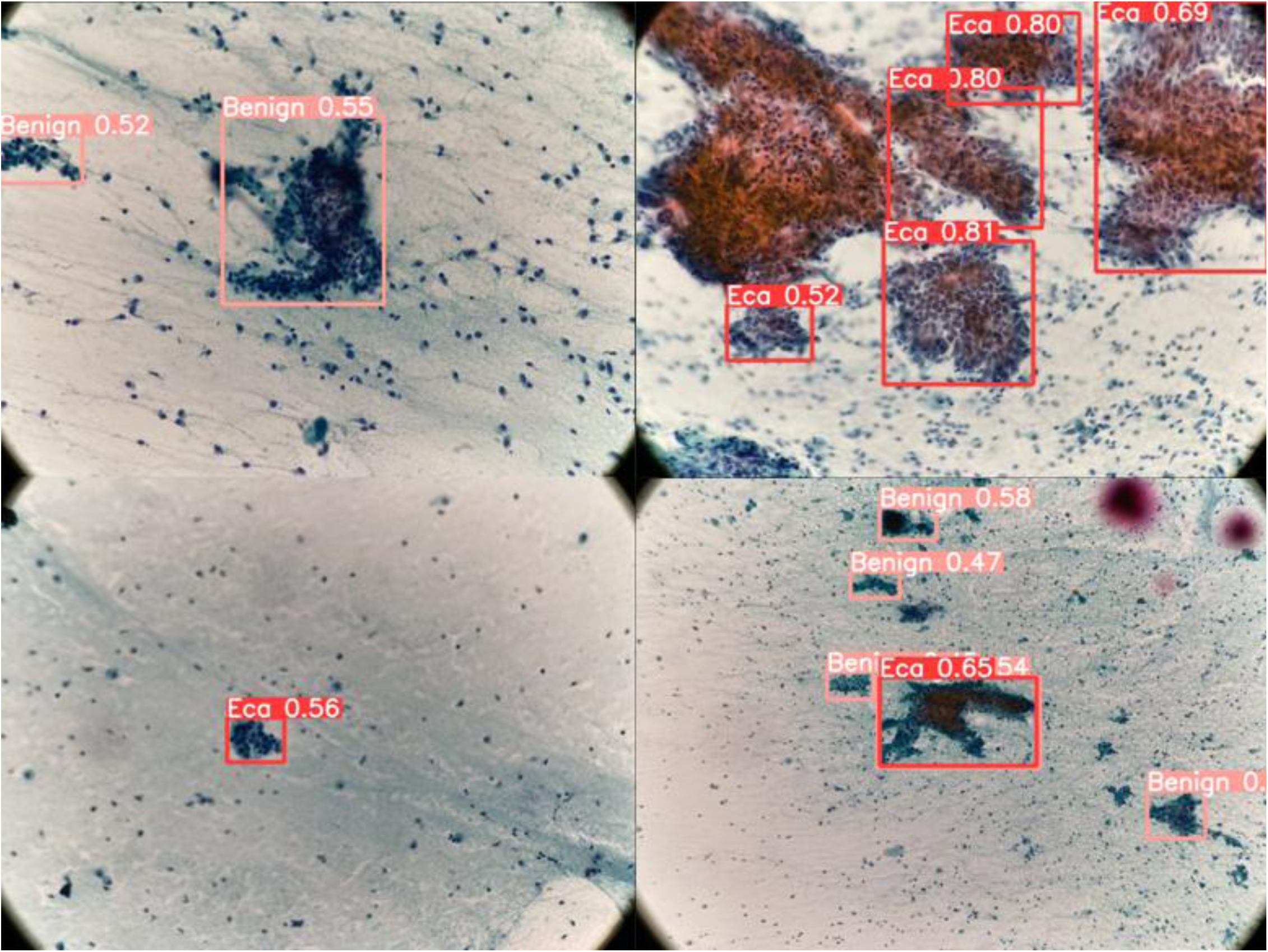
Detection results from the model trained on both benign and malignant cases. The top left shows an image where benign was detected as benign, the top right shows malignant detected as malignant, the bottom left shows benign detected as malignant, and the bottom right shows malignant detected as benign.

### 3.2 Distribution of Perceived Diagnostic Difficulty in AI Model Correct and Incorrect Groups

In this study, 20 images—10 correctly diagnosed by the AI model (Correct Group) and 10 incorrectly diagnosed (Incorrect Group)—were evaluated by three pathologists and eight cytotechnologists.

The median difficulty score for the Correct Group was 2.775, with an interquartile range (IQR) of 0.570, while the Incorrect Group had a median score of 2.910 and an IQR of 0.503. These distributions are shown in the box plot in Figure 4a, explaining how diagnosticians perceived the difficulty of cases that were classified correctly or incorrectly by the AI model.

**Figure 4.**
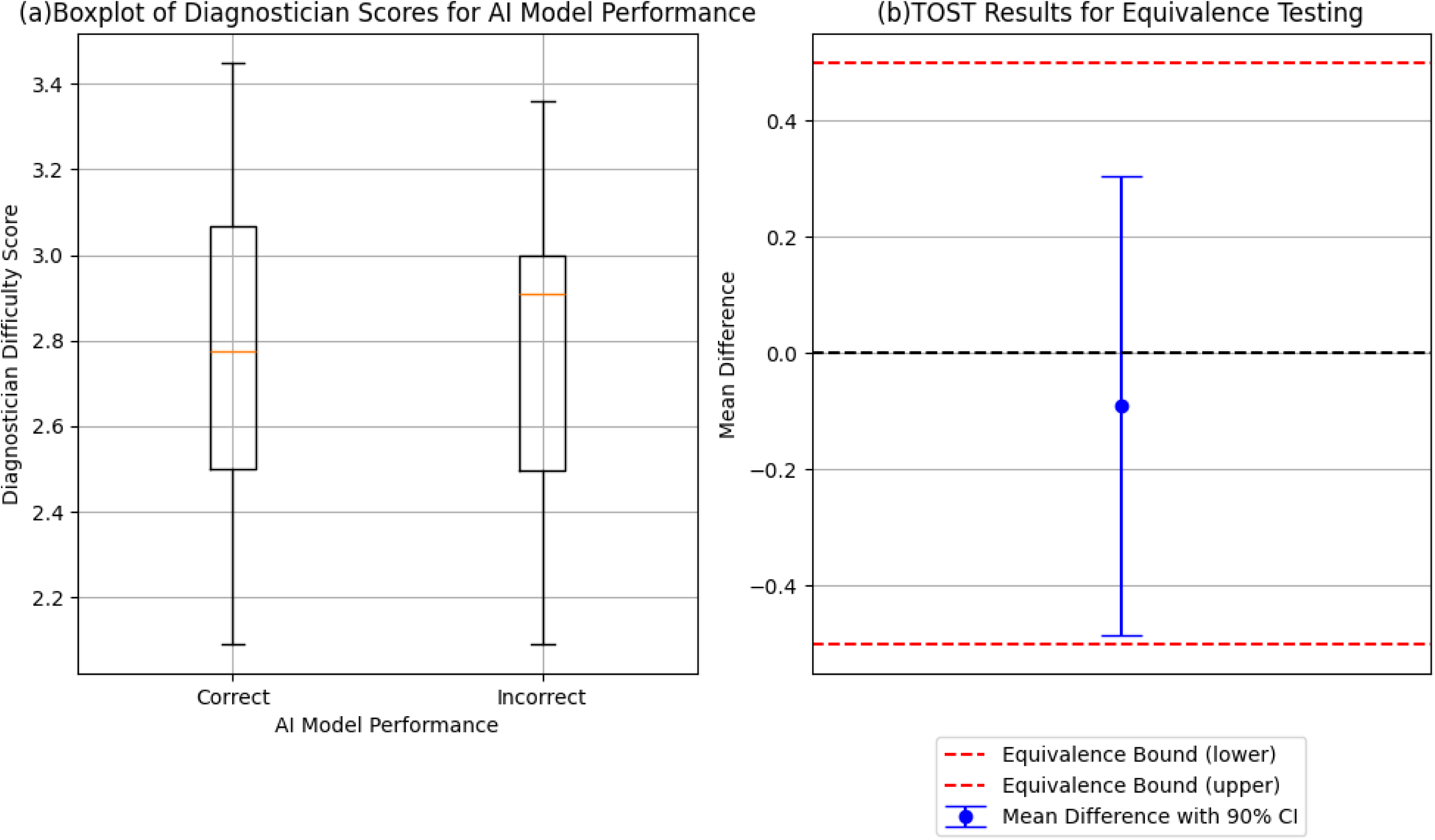
(a) Box Plot of Difficulty Ratings Given by Diagnostic Professionals for Each AI Model Correctness. (b) TOST results showing equivalence between the difficulty assessments of correctly and incorrectly diagnosed images.

### 3.3 Equivalence Test (TOST) for Difficulty Assessment in AI Model Correct and Incorrect Groups

The results of the equivalence test (TOST) showed a lower bound p-value of 0.044, an upper bound p-value of 0.009, and an overall equivalence p-value of 0.044. The mean difference in difficulty scores was -0.090, with a 90% confidence interval of [-0.485, 0.305]. These results indicate that the two groups are statistically equivalent (p < 0.05), as shown in Figure 4b. These statistical tests suggest that there is no significant variation in diagnostic difficulty between the Correct and Incorrect Groups, supporting the potential applicability of the AI model in clinical cytology settings.

### 3.4 Visualization of AI Model Diagnostic Basis Using Grad-CAM

Grad-CAM was used to visualize the AI model’s diagnostic reasoning by highlighting the regions the model focused on during its prediction. The resulting heatmaps represent the level of attention or importance assigned by the AI model, with warmer colors such as red indicating higher relevance and cooler colors like blue indicating lower relevance. This visualization provides cytology specialists and cytotechnologists with insight into the influential features the AI relied upon during diagnosis.

Figure 5 presents examples of Grad-CAM visualizations for both correct and incorrect identifications of benign and malignant clusters. In several cases, the AI model appeared to focus on regions distinct from those typically emphasized by human diagnosticians, for instance, areas lacking prominent nuclear atypia or irregular cluster margins, which are often key features for human interpretation. Instead, the model sometimes extracted features from the central portion of loosely aggregated cell clusters or from peripheral cytoplasmic regions.

**Figure 5.**
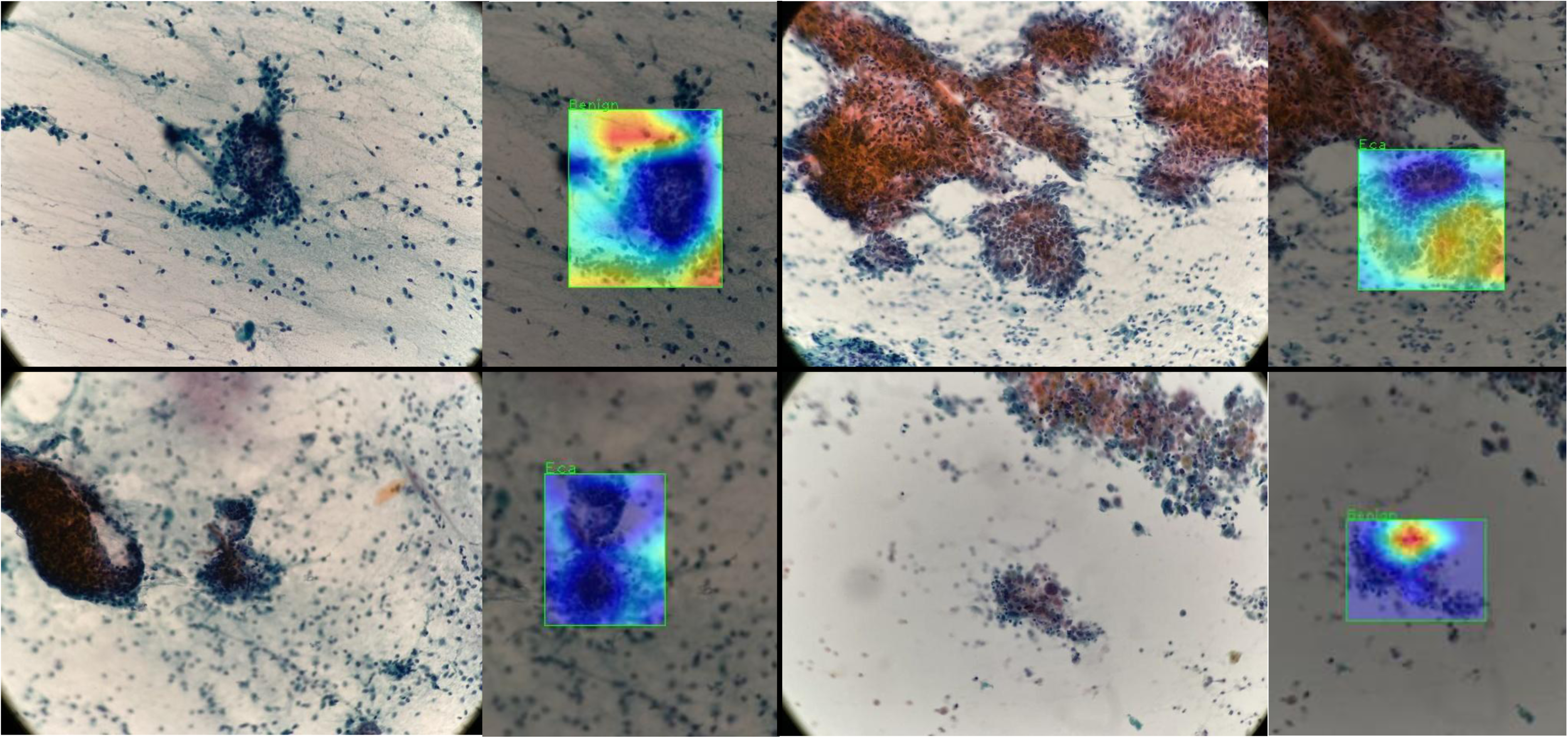
Results of Grad-CAM visualization. The top left image correctly identifies benign cell clusters as benign. The top right image correctly identifies malignant cell clusters as malignant. The bottom left image incorrectly detects benign cell clusters as malignant. The bottom right image incorrectly detects malignant cell clusters as benign.

These observations suggest that AI may be recognizing patterns not conventionally prioritized by experts, raising important considerations regarding the interpretability, reliability, and complementary potential of AI in cytological diagnosis. This discrepancy between AI attention and human reasoning warrants further investigation and is discussed in detail below.

## 4. Discussion

### Summary of Results

In this study, we developed and evaluated AI models focusing on four key objectives: (1) optimizing the annotation strategy, (2) comparing model performance, (3) integrating the AI model into routine diagnostic workflows, and (4) enhancing model explainability and transparency. Our findings address both technical and clinical aspects of endometrial cytology AI implementation.

### 4.2 Annotation of Benign and Malignant Cell Clusters

In histological studies utilizing Whole Slide Imaging (WSI), it is standard practice to annotate both benign and malignant cell clusters.^20^ In contrast, our prior cytological research focused exclusively on annotating malignant clusters, operating under the hypothesis that including benign clusters would introduce noise, thereby adversely affecting the detection of clinically significant malignant cells. To examine this hypothesis, we conducted an experiment comparing both annotation strategies, initially presuming that annotating only malignant clusters would enhance detection accuracy by reducing noise interference.

Contrary to our expectations, the results revealed that annotating both benign and malignant clusters significantly improved the model’s ability to detect malignant cells. Specifically, the YOLOv5x model achieved a mean Average Precision (mAP) of 0.798 when both benign and malignant clusters were annotated, compared to a mAP of 0.769 when only malignant clusters were annotated. These findings underscore the importance of including benign clusters in the annotation process to enhance malignant cell detection accuracy. This insight provides valuable guidance for annotation strategies in AI model development across various pathological domains, particularly in contexts where both benign and malignant clusters coexist or where there may be ambiguity in subtype labeling.

### 4.3 Comparison of YOLOv5x and YOLOv7 Accuracy

To compare the diagnostic performance of YOLOv5x and YOLOv7, hyperparameter tuning was performed under identical conditions in our study, with 100 training epochs and a batch size of 4. As a result, YOLOv5x achieved the highest mAP for both benign and malignant cell clusters, outperforming YOLOv7. No signs of overfitting were observed on the loss curves, suggesting that the inclusion of both benign and malignant annotations contributed to improved generalizability. Interestingly, similar findings have been reported in recent studies involving medical image analysis. The study by Prisilla et al.^21^ demonstrated that YOLOv5x outperformed YOLOv7 in detecting lumbar disc herniation on MRI. While the study by Sadhin et al. ^22^ reported that YOLOv7 showed higher mAP in road damage detection, the YOLOv5 series, including YOLOv5x, still exhibited strong detection precision and utility.

These results suggest that the optimal model architecture may depend heavily on task-specific image characteristics. In cytological images, accurate detection often requires interpreting not only localized features but also the broader context of cell clusters, such as irregular margins, structural overlap, and peripheral cytoplasmic morphology. YOLOv5x, with its larger parameter capacity and expressive power, may be more capable of capturing these global morphological features compared to more compact architectures like YOLOv7. In contrast, YOLOv7’s architectural efficiencies may be better suited to large-scale natural image datasets. These differences highlight the importance of evaluating model suitability within the specific context of clinical applications.

### 4.4 Correlation Between Model and Diagnostician Diagnoses

Previous studies have often examined how AI affects human diagnostic performance, particularly in cases perceived as difficult. For example, in lung cancer detection, high-accuracy AI improved radiologists’ performance in difficult cases, while incorrect AI predictions had adverse effects ^23^. Similarly, the study by Kiani et al ^24^ demonstrated that AI assistance significantly influenced pathologists’ decisions in the histopathologic classification of liver cancer: when the AI model’s prediction was correct, diagnostic accuracy improved, but when incorrect, accuracy decreased—even across different levels of expertise and case difficulty. These studies highlight that the impact of AI in clinical settings is not solely dependent on its accuracy, but also on how its outputs interact with human perception and decision-making.

However, rather than focusing on how AI influences human performance, our study investigates how perceived case difficulty by humans relates to the diagnostic accuracy of the AI model itself. Interestingly, we found no statistically significant difference in perceived difficulty between cases the AI correctly and incorrectly diagnosed. This result suggests that the AI model’s performance was robust regardless of the diagnosticians’ perceived difficulty, and that AI and human experts may be evaluating complexity through fundamentally different frameworks.

Furthermore, recent literature in computational pathology emphasizes a complementary relationship between AI and human diagnosticians. Rather than replacing pathologists, AI is increasingly seen as an assistive tool capable of performing standardized and repetitive tasks with high precision, thereby freeing pathologists to focus on complex cases and integrative decision-making.^25,26^ In particular, AI excels in tasks requiring quantitative consistency, such as biomarker assessment or routine classification, while pathologists provide contextual interpretation based on rare patterns or clinical complexity. This evolving role of AI as a diagnostic assistant aligns with our findings, which suggest that the AI model maintained stable diagnostic accuracy regardless of human-perceived difficulty. Understanding how AI and humans perceive diagnostic complexity differently—and where their assessments diverge—is critical for designing systems that truly integrate into the pathology workflow.

To deepen this understanding, it is essential to visualize how the AI model makes its diagnostic decisions and whether those decisions rely on features similar to those used by human diagnosticians. In our study, we employed Grad-CAM (Gradient-weighted Class Activation Mapping) to visualize the regions the AI model focused on during diagnosis, thereby offering interpretable insights into the model’s reasoning process.

### 4.5 Visualization of AI Diagnostic Basis

To enhance the interpretability of the AI model’s diagnostic decisions, we employed Gradient-weighted Class Activation Mapping (Grad-CAM) to visualize the specific regions within cell clusters that influenced the model’s predictions. Grad-CAM calculates the gradient of the output with respect to the final convolutional layer and combines it with the associated feature maps, producing heatmaps that highlight areas most relevant to the AI’s decision-making process.^15^ This architecture-agnostic technique can be readily applied to convolutional neural networks (CNNs) without requiring structural modifications, making it particularly well-suited for real-time cytological applications.

Grad-CAM has shown promise in various medical imaging domains. For instance, the study by Musthafa et al. ^27^ demonstrated its utility in brain tumor detection using MRI, while Wei et al. ^28^ employed Grad-CAM to visualize prognostic features in colorectal cancer pathology. In cytological diagnosis, where real-time feedback and intuitive interpretation are crucial, Grad-CAM’s ability to directly map influential regions onto diagnostic images offers practical advantages for clinical integration.

However, Grad-CAM does not fully resolve the “black box” issue inherent in deep learning. In our study, there were instances in which the model’s highlighted regions diverged from areas typically emphasized by pathologists—such as nuclear atypia or irregular cluster margins—and instead focused on peripheral cytoplasmic zones or loosely aggregated cell areas. This discrepancy raises the possibility that Grad-CAM visualizations may not always faithfully represent the model’s internal reasoning but rather reflect statistical saliency at the final convolutional layer.

This concern aligns with findings by Saporta et al. ^29^, who reported that Grad-CAM failed to reliably localize diagnostically critical regions in chest X-ray analysis, particularly for pathologies that were small, multifocal, or morphologically complex. Their study also found a positive correlation between model confidence and Grad-CAM reliability, suggesting that saliency maps should be interpreted cautiously in cases where model certainty is low.

In summary, Grad-CAM represents a valuable tool for enhancing transparency in AI-assisted cytological diagnosis, particularly in settings requiring real-time visual feedback. Nevertheless, its limitations—especially the potential misalignment with expert-derived diagnostic features—underscore the need for continued investigation into the fidelity of interpretability tools and their integration into clinical decision-making frameworks.

### 4.6 Limitations

This study has several limitations. First, the dataset size was relatively small, particularly for rare malignant cases, which limits the generalizability of the model. This constraint reflects a broader challenge in cytopathology AI development, where the acquisition of rare cases and accurate annotations is inherently resource-intensive and time-consuming. In addition, the limited availability of publicly accessible annotated medical image datasets presents a major barrier to innovation in computational research and education. Second, all data were collected from a single facility, requiring external validation across multiple institutions and using different imaging equipment. This introduces the risk of domain shift, where a model trained on data from one facility or equipment may perform poorly when applied to data from different settings. Variations in imaging conditions, such as magnification, lighting, and even patient demographics, can significantly impact the model’s ability to generalize. Therefore, it is crucial to test the model across diverse institutions and imaging modalities to ensure broader applicability.

Additionally, as discussed with Grad-CAM, the diagnostic basis visualized by the method was unclear in some cases, further highlighting unresolved aspects of the AI model’s black-box nature. Lastly, the model’s performance is dependent on hardware constraints and computational resources, which may limit its scalability in clinical practice. Addressing these limitations will require larger datasets, collaboration with multiple institutions, and continued development of methods to improve both explainability and robustness. In particular, non-visual XAI methods—such as auxiliary explanations, case-based explanations, and textual explanations—may serve as promising alternatives or complements to visual techniques like Grad-CAM in resolving the black-box challenges of deep learning models ^30^.

### 4.7 Significance of the Developed Model

Although this study focuses on cell clusters, slide-level performance is crucial for clinical use. Our AI model integrates with a microscope and a CCD camera, allowing diagnosticians to observe AI-generated judgments and explanations in real time on a nearby screen. While 30 frames per second (FPS) is generally considered sufficient for real-time detection, our previous research has demonstrated that our model can achieve 60 FPS. This higher frame rate ensures smooth operation, minimizing any potential delays or stress for diagnosticians as they move the slide. The ability to provide real-time feedback at this speed enhances the clinical workflow by reducing diagnostic time and promoting workstyle reform in medical settings, such as decreasing labor hours and improving overall efficiency. Furthermore, many medical AI research papers focus solely on accuracy metrics, and few prior studies have investigated the correlation between AI model performance and the assessments made by the diagnosticians who intend to use these models, as this study does. By incorporating this perspective, our work not only contributes to technical advancement but also aligns with the practical realities of clinical deployment. Moreover, in the development of medical AI models, many healthcare professionals hold the preconception that AI might threaten their job security ^31^, necessitating development approaches that are more aligned with the needs of healthcare providers. This study proposes a new direction for the practical implementation of medical AI models by evaluating the correlation between diagnosticians’ perceived diagnostic difficulty and the AI model’s accuracy, ensuring that the model is developed not only with consideration for the diagnosticians’ role and perspective but also as a tool to support and enhance their diagnostic capabilities.

Furthermore, the challenges encountered in this study—such as acquiring rare cases, annotating cytological images, and ensuring real-time compatibility—are not unique to endometrial cytology. They reflect broader issues in implementing AI in pathology. Therefore, our strategy, including microscope-integrated inference and explainability visualization, may be extended to other cytological and pathological domains, such as the analysis of bone marrow aspirates ^32^. where real-time decision support and interpretable results are equally critical. Ultimately, we envision that this model will serve as a foundation for broader applications, such as automated screening support in resource-limited settings or educational tools for training cytotechnologists and pathologists, further expanding the utility of AI in medicine.

## 5. Conclusion

In this study, we developed and evaluated AI models based on YOLOv5x and YOLOv7, with YOLOv5x achieving the highest diagnostic accuracy (mAP 0.798) for both benign and malignant cell cluster annotations. The annotation strategy that targeted both benign and malignant clusters was a novel aspect in the field of pathology AI. The investigation into the correlation between AI model performance and perceived diagnostic difficulty by cytology professionals revealed that the AI maintained consistent accuracy, regardless of case complexity. Additionally, the use of Grad-CAM for visualizing diagnostic reasoning significantly enhanced the transparency and interpretability of the model, making it more reliable for clinical application. These advancements highlight the potential of integrating AI-assisted tools into pathological workflows to improve diagnostic precision and efficiency.

The high-performing YOLOv5x model, its robust performance across varying levels of case difficulty, and the enhanced model interpretability through Grad-CAM visualization emphasize the importance of AI-human collaboration in medical diagnosis. This study represents a significant milestone in the development of AI-assisted pathology tools, offering valuable insights for future research and clinical use. Furthermore, the focus on model transparency and consistent performance strengthens the role of AI in healthcare, ultimately contributing to more accurate and reliable diagnostic outcomes. Importantly, because this system operates without the need for whole slide scanners and integrates directly with standard microscopes, it holds potential for broader applications beyond endometrial cytology, including other cytological and pathological workflows.

## Data Availability

All data produced in the present study are available upon reasonable request to the authors

## Author contributions

I.S. and M.T. performed study concept and design, sample collection, development of methodology, and writing, review, and revision of the paper; S. Tanaka performed development of methodology, and writing, review, and revision of the paper; E.T. and Y.T. provided acquisition and statistical analysis of data, and review and revision of the paper. S. Takakuma, Y.K., S.H and S.K. provided technical and material support; Y.T., and A.S. reviewed the study concept and design, as well as the paper. All the authors read and approved the final manuscript. We would also like to express our sincere gratitude to the cytopathologists Y. Murase, Y. Watarai, H. Kamaguchi, Y. Kimura, M. Aida, S. Suzuki, N. Inoue, and S. Nagai for their invaluable cooperation in this study.

## Acknowledgments

We extend our gratitude to Arimi Ishikawa, Naomi Kuwahara, and Masaya Okaizumi for their technical support throughout this study. We also thank Kazuhiro Takeuchi, Hideaki Kuno, and Emi Sakamoto for their contributions to the education of medical students, and Kenta Tominaga for his unwavering support. Furthermore, we appreciate the development of YOLOv5x by Ultralytics and Mohammadi Kazaj, Pooya, who connects YOLOv5x and Grad-CAM.

