## Supplementary Figure S1 for "Development and Clinical Application of a Deep Learning-Based Endometrial Cancer Cytology Supporting Model"

### Slide 1
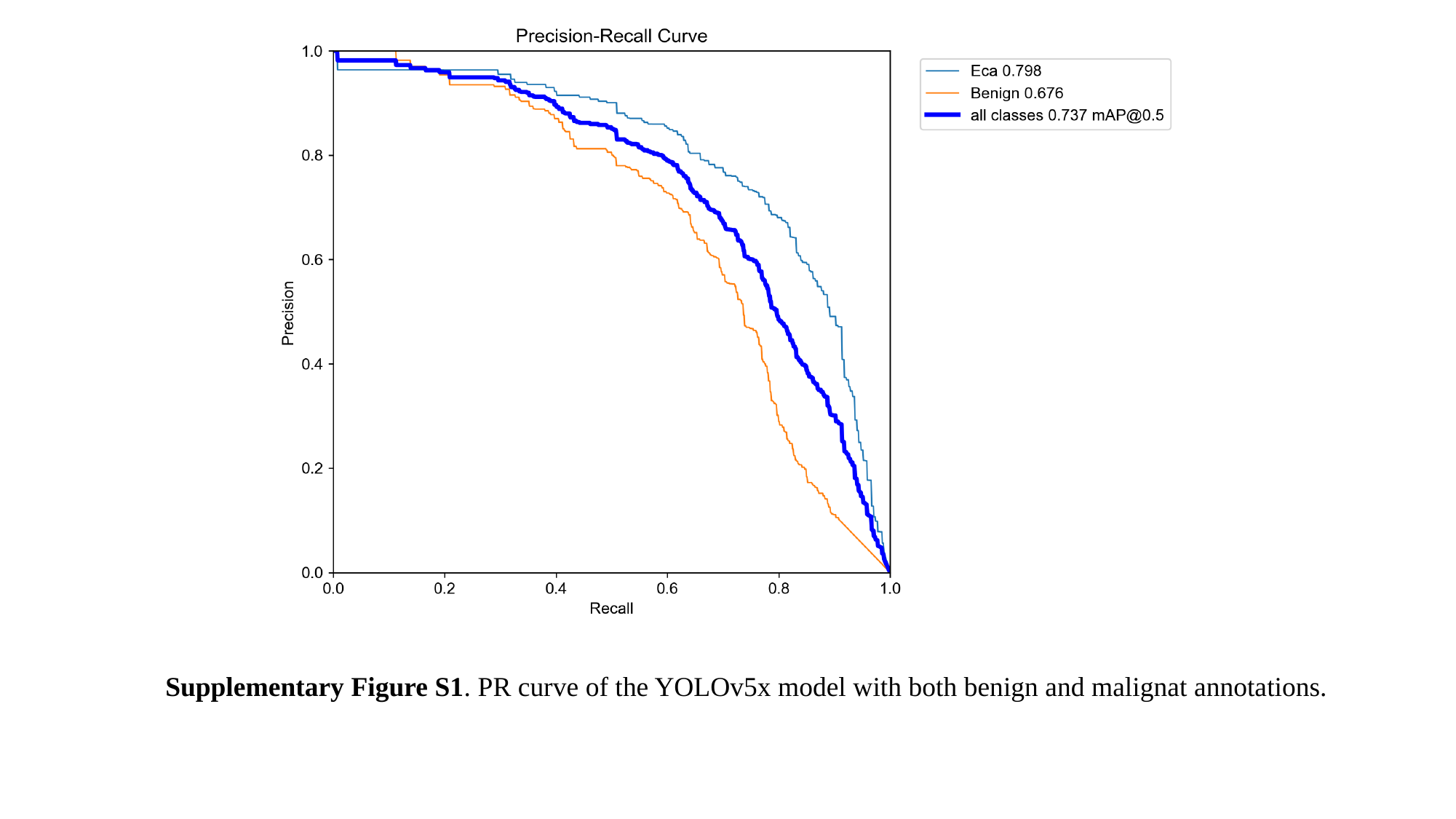

Supplementary Figure S1. PR curve of the YOLOv5x model with both benign and malignat annotations.
