## Supplementary Figure S2 for "Development and Clinical Application of a Deep Learning-Based Endometrial Cancer Cytology Supporting Model"

### Slide 1
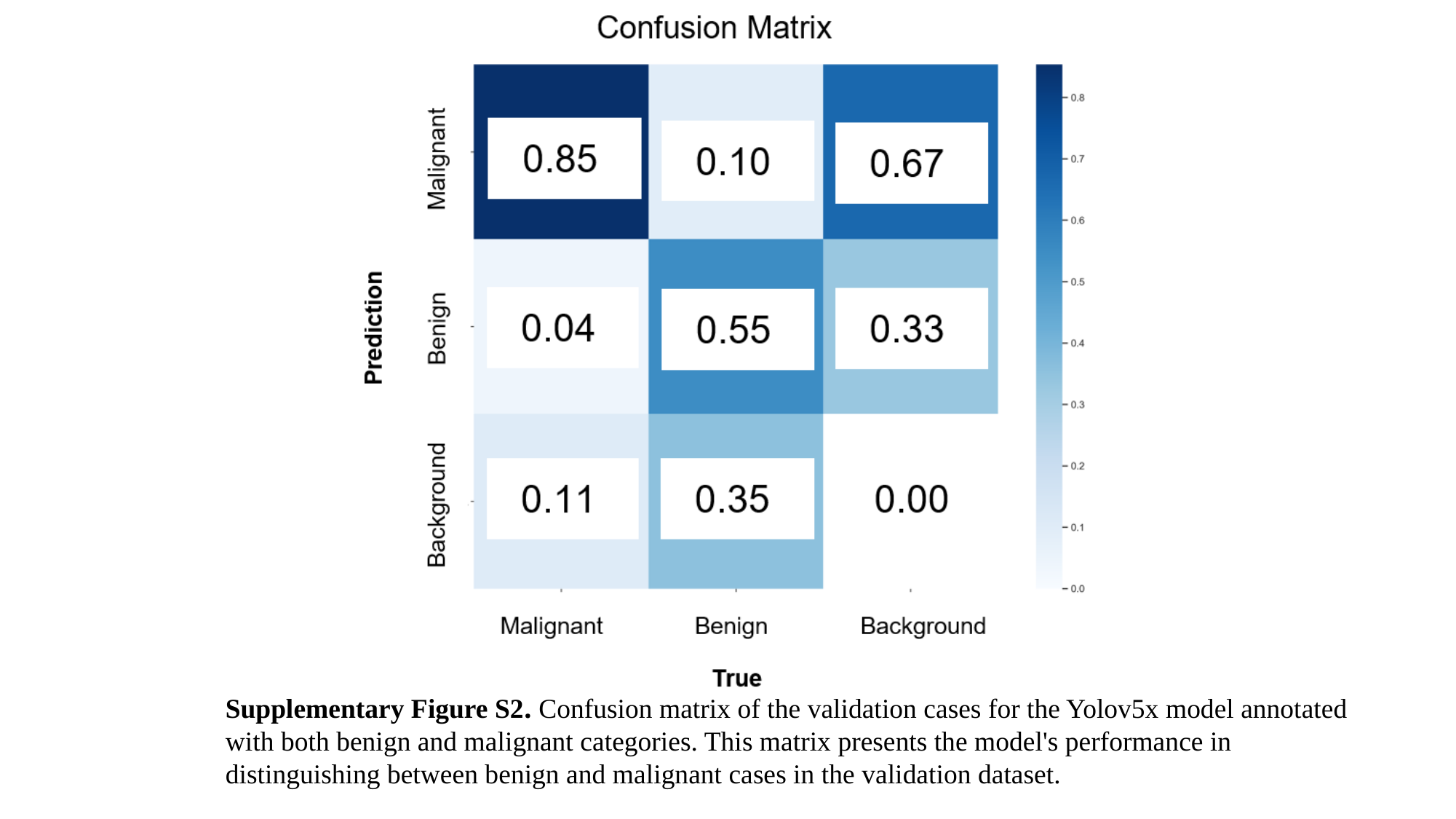

Supplementary Figure S2. Confusion matrix of the validation cases for the Yolov5x model annotated with both benign and malignant categories. This matrix presents the model's performance in distinguishing between benign and malignant cases in the validation dataset.
